# ETIOLOGY OF PRIMARY CEREBELLAR INTRACEREBRAL HEMORRHAGE BASED ON TOPOGRAPHIC LOCALIZATION

**DOI:** 10.1101/2023.06.20.23291683

**Authors:** Diego Incontri, Sarah Marchina, Alexander Andreev, Mitchell Wilson, Jia-Yi Wang, David Lin, Elizabeth C. Heistand, Filipa Carvalho, Magdy Selim, Vasileios-Arsenios Lioutas

**Author notes:** **Corresponding author:** Diego Incontri, MD, Email address, Address: Department of Neurology, Stroke Division, Beth Israel Deaconess Medical Center and Harvard Medical School, 330 Brookline Avenue, Boston, MA 02215, USA.

## Abstract

**Background:** Cerebellar intracerebral hemorrhage (cICH) is often attributed to hypertension or cerebral amyloid angiopathy (CAA). However, deciphering the exact etiology can be challenging. A recent study reported a topographic etiologic relationship with superficial cICH secondary to CAA. We aimed to re-examine this relationship between topography and etiology in a separate cohort of patients and using the most recent Boston criteria version 2.0.

**Methods:** Retrospective analysis of consecutive patients with primary cICH admitted to a tertiary academic center between 2000-2022. cICH location on brain CT/MRI scan(s) was divided into strictly superficial (cortex, surrounding white matter, vermis), vs deep (cerebellar nuclei, deep white matter) or mixed (both regions). MRIs were rated for markers of cerebral small vessel disease. We assigned possible/probable vs absent CAA using Boston criteria 2.0.

**Results:** We included 197 patients; 106 (53.8%) females, median age 74 (63-82) years. Fifty-six (28%) patients had superficial cICH and 141 (72%) deep/mixed cICH. MRI was available for 112 (57%) patients [30(26.8%) with superficial and 82(73.2%) with deep/mixed cICH]. Superficial cICH patients were more likely to have possible/probable CAA (48.3% vs 8.6%; OR 11.43, 95%CI 3.26-40.05; p<0.001), strictly lobar cerebral microbleeds (CMBs) (51.7% vs 6.2%; OR 14.18, 95%CI 3.98-50.50; p<0.001), and cortical superficial siderosis (13.8% vs 1.2%; OR 7.70, 95%CI 0.73-80.49; p=0.08). Patients with deep/mixed cICH patients were more likely to have deep/mixed CMBs (59.2% vs 3.4%; OR 41.39, 95% CI 5.01-341.68; p=0.001), lacunes (54.9% vs 17.2%; OR 6.14, 95% CI 1.89-19.91; p=0.002), severe basal ganglia enlarged perivascular spaces (36.6% vs 7.1%; OR 7.63, 95% CI 1.58-36.73; p=0.01), hypertension (84.4% vs 62.5%; OR 3.43, 95% CI 1.61,-7.30; p=0.001), and higher admission systolic blood pressure (172 [146-200] vs 146 [124-158] mmHg, p<0.001).

**Conclusions:** Our results suggest that superficial cICH is strongly associated with CAA whereas deep/mixed cICH is almost exclusively linked to hypertensive arteriopathy.

## Introduction

Spontaneous ICH accounts for 10-15% of all strokes,^1^ with the subgroup of patients with cerebellar ICH (cICH) accounting for approximately 10% of all ICHs and 1.5% of all strokes.^2^

In adults over 55 years of age, cerebral small vessel disease (cSVD) is the most common cause of spontaneous ICH.^3^ cSVD presents with two main etiologic subtypes: 1) arteriolosclerosis due to hypertensive arteriopathy; and 2) cerebral amyloid angiopathy (CAA).^4^ These two underlying processes are associated with distinct topographic localization of ICH^5-7^ and distribution patterns of cSVD markers.^8-13^ Patients with a lobar ICH and strictly lobar cSVD markers are more likely to have CAA as the predominant microangiopathy, while those with a deep ICH (basal ganglia, thalamus, deep white matter, and pons) and predominantly deep cSVD markers are more likely to have arteriolosclerosis as the predominant microangiopathy.

While the above associations are relatively well-established for supratentorial and brainstem primary ICH, the understanding of the etiology and underlying pathology of spontaneous cICH, as well as the significance of its topographic localization, is less well-established. Previous studies have linked spontaneous cICH to hypertension,^14, 15^ particularly when it occurs near the cerebellar nuclei and surrounding deep white matter.^16^ Studies exploring pathological and radiological findings of CAA did not examine cerebellar involvement^17^ and have not capitalized on recent advances in neuroimaging markers of cSVD. Neuropathological studies suggest that a subgroup of cICH might be related to CAA, particularly when located in the superficial region of the cerebellum, including the cerebellar cortex and vermis.^18, 19^ Additionally, recent radiological studies have indicated that superficial cICH is associated with magnetic resonance imaging (MRI) markers of CAA, such as strictly lobar cerebral microbleeds.^20^ Therefore, the purpose of this study was twofold: to validate previous results in an external cohort and to use MRI markers of cSVD to determine whether superficial cICH is associated with CAA according to the most recent version of the Boston criteria 2.0.

## Methods

We performed a retrospective analysis of consecutive patients with primary cICH admitted to a tertiary academic center between October 2000 and February 2022. We excluded patients with a secondary cause of cICH, such as those with antecedent head trauma, hemorrhagic transformation of ischemic infarct, arteriovenous malformation, cavernoma, hemorrhagic tumor/metastasis, and central nervous system vasculitis. We collected the following baseline characteristics: age, gender, vascular risk factors, blood pressure and serum glucose on presentation, the National Institutes of Health Stroke Scale (NIHSS) score, ICH score,^21^ presence of intraventricular hemorrhage (IVH); and anticoagulation, antiplatelet or statin use. The study was approved by the local institutional review board and ethics committee.

### Imaging assessments and rating

Admission computed tomography (CT) scan and/or MRI were used to categorize cICH location into superficial (cerebellar cortex, surrounding white matter, and vermis), deep (cerebellar nuclei and deep white matter), or mixed (both regions) (*Figure 1 & 2*). We divided the patients into two groups: 1) deep/mixed cICH; and 2) superficial cICH.

**Figure 1.**
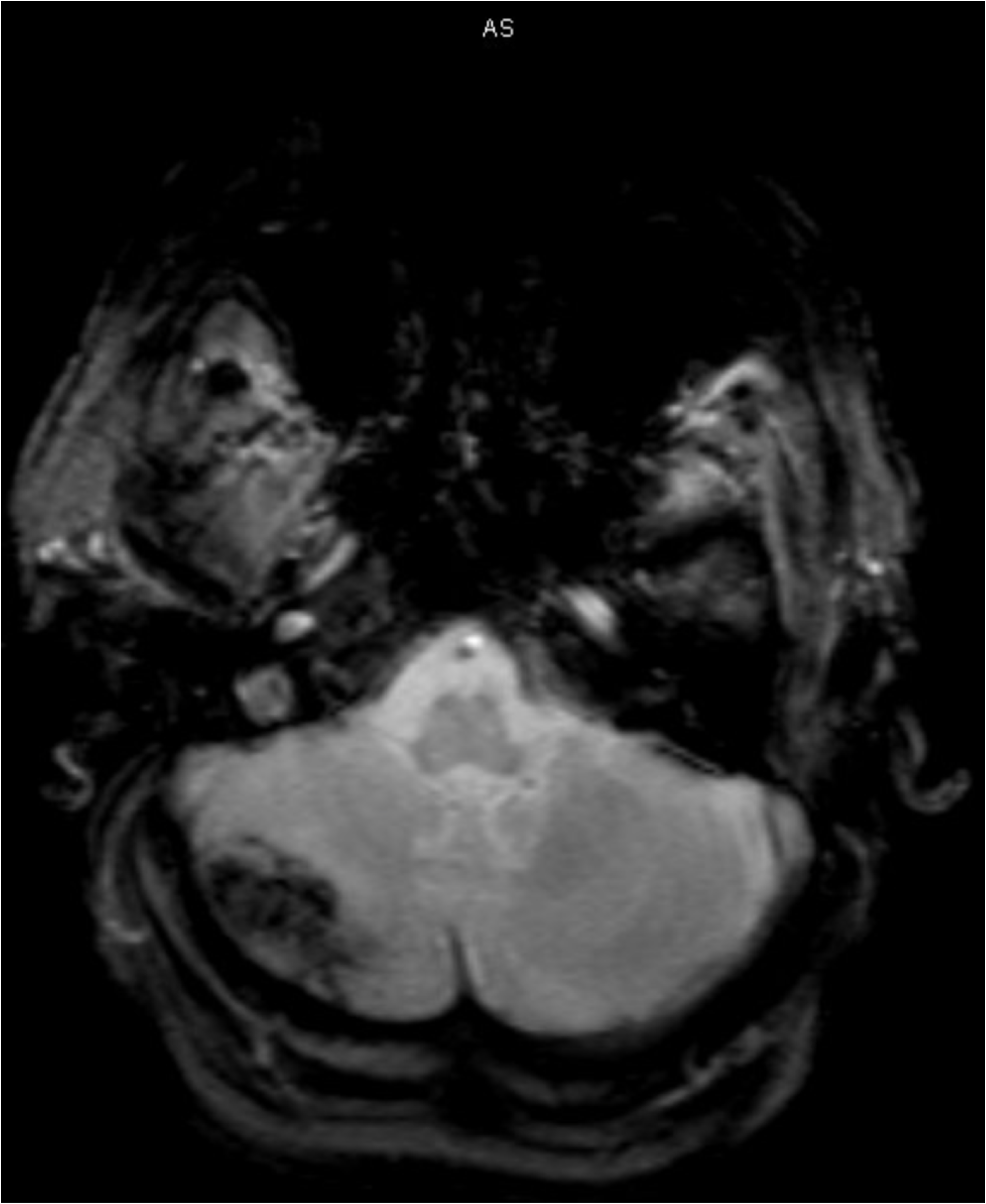
Superficial cerebellar hemorrhage on T2*-weighted gradient-recalled echo (GRE), representative image. The superficial region of the cerebellum corresponds to the cerebellar cortex, surrounding white matter, and vermis.

**Figure 2.**
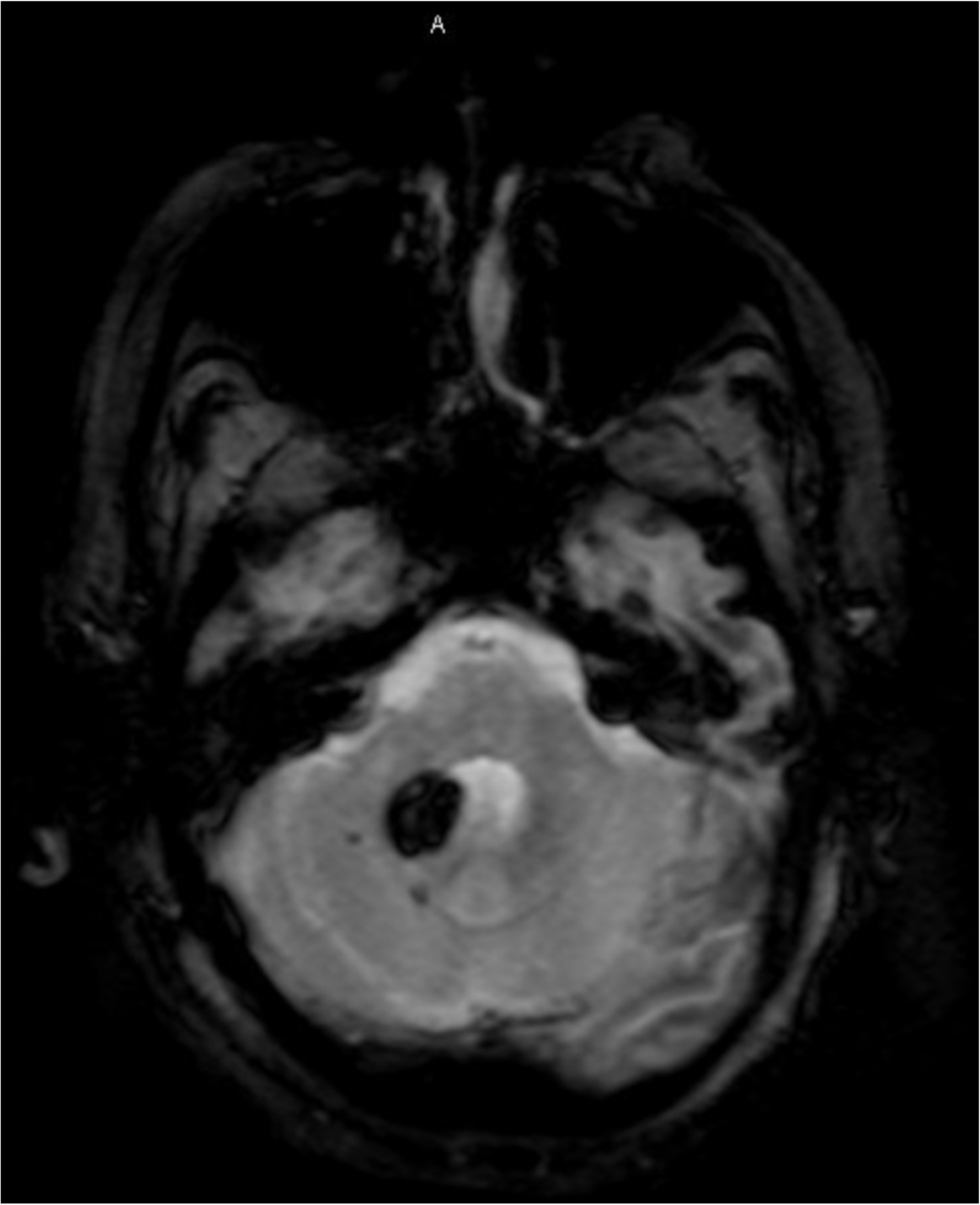
Deep cerebellar hemorrhage on T2*-weighted gradient-recalled echo (GRE), representative image. The deep region of the cerebellum corresponds to the cerebellar nuclei and deep white matter.

In those patients who had an MRI done during the admission period, cSVD markers were rated according to the STRIVE consensus.^22^ Specifically, we rated MRIs for supratentorial cerebral microbleeds (CMBs), cortical superficial siderosis (cSS), lacunes, enlarged perivascular spaces (EPVS) in both the basal ganglia and centrum semiovale, both periventricular and deep white matter hyperintensities (WMH) using the Fazekas score, and WMH in a multispot pattern. Supratentorial CMBs were categorized based on their distribution into strictly lobar or strictly deep/mixed.^20^ EPVS were dichotomized into severe (score > 20) or non-severe (score ≤ 20).^23^ The multispot WMH pattern was defined as the presence of more than 10 small circular or ovoid T2-weighted FLAIR hyperintense lesions in the bilateral subcortical white matter.^24^

The cSVD rating was conducted by 3 raters (DI, SM, and FC) prior to the commencement of this project as part of our prospective ICH registry which includes all ICH (both supra- and infratentorial) and preceded the present project conception by several months. Interrater agreement for various markers was assessed on a sample of 30 randomly selected brain MRIs. Cronbach’s alpha was 0.79 for presence of CMBs, 0.73 for presence of lacunes, 0.87 for EPVS in basal ganglia rating, 0.73 for EPVS in centrum semiovale rating, 0.87 for deep WMH Fazekas score, and 0.86 for periventricular WMH Fazekas score. cICH localization was performed by 2 raters, VL and DI, both blinded to the cSVD rating.

Based on the most recent Boston criteria version 2.0,^25^ patients aged ≥ 50 years presenting with spontaneous ICH (cICH for this study) and at least two of the following strictly lobar T2*-weighted MRI markers were assigned a diagnosis of probable CAA: ICH, CMBs, cSS or convexity subarachnoid hemorrhage. If only one of these latter MRI markers was present, and in the absence of a secondary cause of ICH, a diagnosis of possible CAA was made. To avoid self-selection of patients who met the CAA criteria, we did not use the second section of the probable and possible CAA criteria, as cICH was not counted as a lobar or deep hemorrhagic lesion in this section. Finally, based on this most recent version of the Boston criteria, we compared the association between deep/mixed cICH and superficial cICH with the diagnosis of possible or probable CAA. The diagnoses of definite or probable CAA with supporting pathology were not given due to lack of pathology samples.

### Statistical analysis

We used the Chi-square and Fisher’s exact test to compare demographic, clinical, and neuroimaging characteristics between deep/mixed cICH and superficial cICH. Categorical variables were reported as absolute counts and percentages of the total available data. Continuous variables were reported as means (± standard deviation) or median (interquartile range). We used the Student’s t-test for normally distributed continuous variables and the Mann–Whitney U test for not normally distributed continuous variables. The Shapiro–Wilk test was used to assess the normality of the distribution for continuous variables.

We first examined univariable associations between predictive variables of interest (cSVD markers) and our outcome of interest (cICH location). Subsequently, age, gender, and variables with a p value < 0.1 in univariate analysis were entered in a multivariable logistic regression model. We report Odds Ratios and 95% confidence intervals. Bonferroni correction was used, given multiple testing, and the significance level was determined at a 2-sided p-value of 0.005. All analyses were performed using STATA.17 (StataCorp, College Station, TX, USA).

## Results

We identified 197 patients with primary cICH. The median age was 74 (63-82) years, and 106 (53.8%) patients were females. One hundred and forty-one (72%) patients had deep/mixed and 56 (28%) had superficial cICH. *Table 1* shows the differences in baseline characteristics between both groups. Patients with deep/mixed cICH were more likely to have a history of HTN (84.4% vs 62.5%, p=0.001) (OR 3.43, 95% CI 1.61,-7.30; p=0.001), prior ischemic stroke (22% vs 8.9%, p=0.03), presence of IVH (39.7% vs 17.9%, p=0.003), and higher systolic blood pressure (172 [146-200] vs 146 [124-158] mmHg, p<0.001) and mean arterial pressure (113.3 [99.3-130.7] vs 97 [86-114] mmHg, p<0.001) on admission when compared to those with superficial cICH. Patients with superficial cICH were more likely to have a medical history of coronary artery disease when compared to those with deep/mixed cICH (30.4% vs 13.5%, p=0.006). We did not find a statistically significant difference in the use of antiplatelet agents (38.2% vs 34.8, p=0.65) or anticoagulants (40% vs 32.9, p=0.34) between patients with superficial or deep/mixed cICH.

**Table 1.**
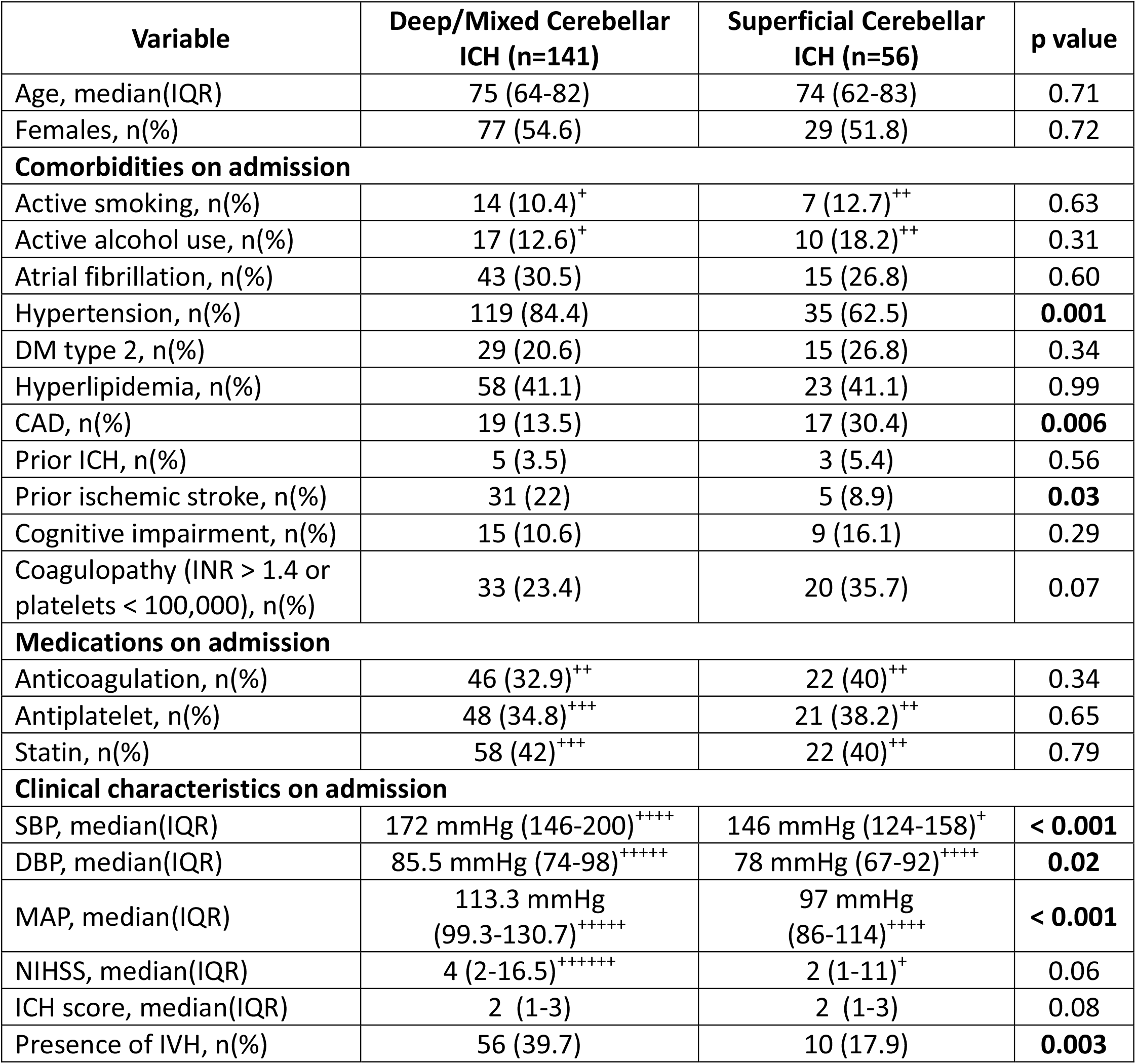
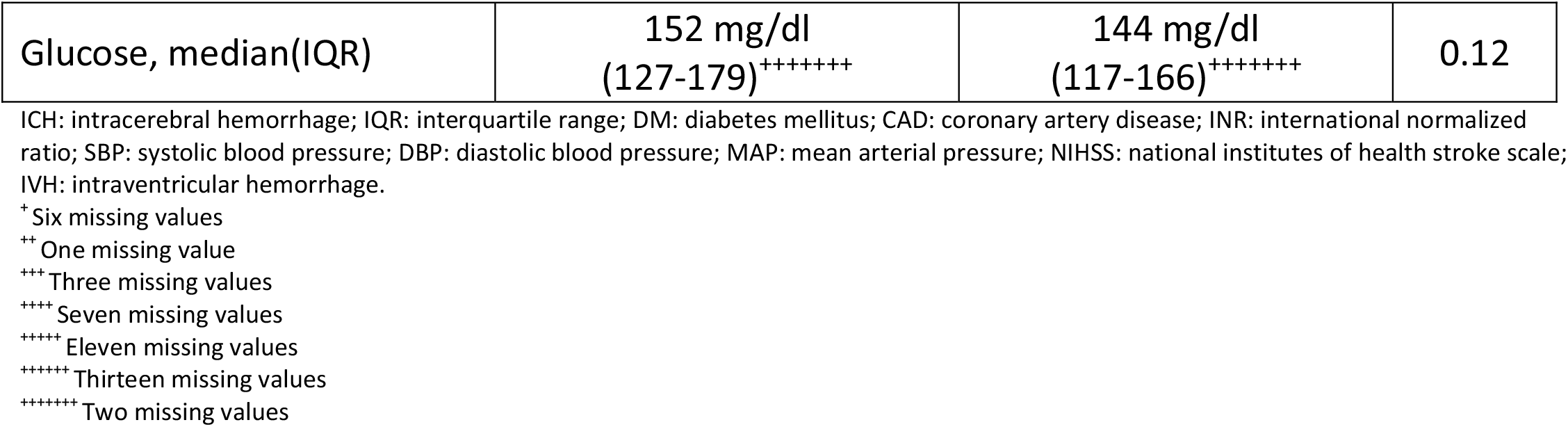
Differences in baseline characteristics between patients with deep/mixed cerebellar ICH and superficial cerebellar ICH.

Of the 197 patients with primary cICH, 112 underwent MRI during admission. Of those, 82 (73.2%) had deep/mixed, and 30 (26.8%) had superficial cICH. *Table 2* shows the differences in MRI markers of cSVD between both groups. Univariate analysis showed that patients with superficial cICH were more likely to have a diagnosis of possible or probable CAA according to the Boston criteria version 2.0 (48.3% vs 8.6%, p< 0.001). In addition, patients with superficial cICH were more likely to have strictly lobar CMBs (51.7% vs 6.2%, p<0.001), cSS (13.8% vs 1.2%, p=0.02), and WMH in a multispot pattern (30% vs 12.4%, p=0.03) when compared to those with deep/mixed cICH. On the other hand, patients with deep/mixed cICH were more likely to have deep/mixed CMBs (59.2% vs 3.4%, p<0.001), lacunes (54.9% vs 17.2%, p=0.001), and severe EPVS in the basal ganglia (36.6% vs 7.1%, p=0.005).

**Table 2.** Differences in MRI markers of cSVD between patients with deep/mixed cerebellar ICH and superficial cerebellar ICH.

| Variable | Deep/Mixed Cerebellar ICH (n=82) | Superficial Cerebellar ICH (n=30) | OR (95% CI)* | p value |
| --- | --- | --- | --- | --- |
| Possible or probable CAA, n(%)** | 7 (8.6)<sup>+</sup> | 14 (48.3)<sup>+</sup> | 9.86<br>(3.40-28.58) | < 0.001 |
| Lobar CMBs, n(%) | 5 (6.2)<sup>+</sup> | 15 (51.7)<sup>+</sup> | 16.28<br>(5.09-52.03) | < 0.001 |
| Deep/Mixed CMBs, n(%) | 48 (59.2)<sup>+</sup> | 1 (3.4)<sup>+</sup> | 40.72<br>(5.27-314.25) | < 0.001 |
| cSS, n(%) | 1 (1.2)<sup>+</sup> | 4 (13.8)<sup>+</sup> | 12.8<br>(1.36-119.85) | 0.02 |
| Presence of lacunes, n(%) | 45 (54.9) | 5 (17.2)<sup>+</sup> | 5.83<br>(2.02-16.80) | 0.001 |
| Severe EPVS in BG, n(%) | 26 (36.6)<sup>+++</sup> | 2 (7.1)<sup>++</sup> | 8.71<br>(1.93-39.29) | 0.005 |
| Severe EPVS in CSO, n(%) | 19 (26.8)<sup>+++</sup> | 11 (39.3)<sup>++</sup> | 1.77<br>(0.70-4.45) | 0.22 |
| PV WMH Fazekas 2-3, n(%) | 65 (80.2)<sup>+</sup> | 20 (66.7) | 2.03<br>(0.79-5.17) | 0.13 |
| Deep WMH Fazekas 2-3, n(%) | 45 (55.6)<sup>+</sup> | 13 (43.33) | 1.63<br>(0.70-3.80) | 0.25 |
| Multispot WMH pattern, n(%) | 10 (12.4)<sup>+</sup> | 9 (30) | 3.04<br>(1.09-8.46) | 0.03 |

MRI: magnetic resonance imaging; cSVD: cerebral small vessel disease; ICH: intracerebral hemorrhage; OR: odds ratio; CI: confidence interval; CAA: cerebral amyloid angiopathy; CMBs: cerebral microbleeds; cSS: cortical superficial siderosis; EPVS: enlarged perivascular spaces; BG: basal ganglia; CSO: centrum semiovale; PV: periventricular; WMH: white matter hyperintensities.
\*Unadjusted analysis
\*\*By Boston criteria version 2.0
\* One missing value
++ Two missing values
+++ Eleven missing values

In the multivariable logistic regression model (*Table 3*), presence of strictly lobar CMBs (OR 14.18, 95% CI 3.98-50.50; p<0.001) and a diagnosis of possible or probable CAA (OR 11.43, 95% CI 3.26-40.05; p<0.001) were associated with superficial cICH. Conversely, presence of deep/mixed CMBs (OR 41.39, 95% CI 5.01-341.68; p=0.001) or lacunes (OR 6.14, 95% CI 1.89-19.91; p=0.002), were associated with deep/mixed cICH. The OR for severe EPVS in the basal ganglia was 7.63 (95% CI 1.58-36.73), but did not reach statistical significance after Bonferroni correction (p=0.01).

**Table 3.** Multivariable adjusted analysis exploring the association between MRI markers of cSVD and deep/mixed cerebellar ICH or superficial cerebellar ICH.

| Variable | Deep/Mixed Cerebellar ICH (n=82) | Superficial Cerebellar ICH (n=30) | OR (95% CI)* | p value |
| --- | --- | --- | --- | --- |
| Possible or probable CAA, n(%)** | 7 (8.6)<sup>+</sup> | 14 (48.3)<sup>+</sup> | 11.43 (3.26-40.05) | < 0.001 |
| Lobar CMBs, n(%) | 5 (6.2)<sup>+</sup> | 15 (51.7)<sup>+</sup> | 14.18 (3.98-50.50) | < 0.001 |
| Deep/Mixed CMBs, n(%) | 48 (59.2)<sup>+</sup> | 1 (3.4)<sup>+</sup> | 41.39 (5.01-341.68) | 0.001 |
| cSS, n(%) | 1 (1.2)<sup>+</sup> | 4 (13.8)<sup>+</sup> | 7.70 (0.73-80.49) | 0.08 |
| Presence of lacunes, n(%) | 45 (54.9) | 5 (17.2)<sup>+</sup> | 6.14 (1.89-19.91) | 0.002 |
| Severe EPVS in BG, n(%) | 26 (36.6)<sup>+++</sup> | 2 (7.1)<sup>++</sup> | 7.63 (1.58-36.73) | 0.01 |
| Severe EPVS in CSO, n(%) | 19 (26.8)<sup>+++</sup> | 11 (39.3)<sup>++</sup> | 1.78 (0.63-4.97) | 0.26 |
| PV WMH Fazekas 2-3, n(%) | 65 (80.2)<sup>+</sup> | 20 (66.7) | 1.77 (0.58-5.39) | 0.31 |
| Deep WMH Fazekas 2-3, n(%) | 45 (55.6)<sup>+</sup> | 13 (43.33) | 1.50 (0.57-3.98) | 0.40 |
| Multispot WMH pattern, n(%) | 10 (12.4)<sup>+</sup> | 9 (30) | 2.19 (0.71-6.71) | 0.17 |

MRI: magnetic resonance imaging; cSVD: cerebral small vessel disease; ICH: intracerebral hemorrhage; OR: odds ratio; CI: confidence interval; CAA: cerebral amyloid angiopathy; CMBs: cerebral microbleeds; cSS: cortical superficial siderosis; EPVS: enlarged perivascular spaces; BG: basal ganglia; CSO: centrum semiovale; PV: periventricular; WMH: white matter hyperintensities.
\*Multivariable adjusted analysis
\*\*By Boston criteria version 2.0
<sup>+</sup> One missing value
<sup>++</sup> Two missing values
<sup>+++</sup> Eleven missing values

## Discussion

We found that superficial cICH localization is associated with higher prevalence of MRI markers of CAA, such as strictly lobar CMBs, cSS, and WMH in a multispot pattern, and possible/probable CAA based on the Boston criteria 2.0. Conversely, deep/mixed cICH was associated with history of hypertension, prior ischemic stroke, higher systolic blood pressure and mean arterial pressure on admission, and MRI markers of hypertensive arteriopathy, such as deep/mixed CMBs, lacunes, and severe EPVS in the basal ganglia.

Our present work is in agreement with prior studies^15, 20, 23^ suggesting that the topographic localization of the hemorrhage within the cerebellum mirrors that of the cerebral cortex and subcortical regions, and signifies different pathophysiological mechanisms of cSVD. Interestingly, our findings correlate with the study of Duvernoy et al.,^26^ indicating that intracortical cerebellar vessels closely resemble those of the cerebral cortex; and that vascular amyloid deposition involving the cerebellum is usually confined to cerebellar cortical and leptomeningeal vessels,^18, 27, 28^ a finding analogous to what occurs in the supratentorial region. Conversely, our study also showed that patients with deep/mixed cICH are more likely to have hypertensive arteriopathy as the primary etiology. This finding is likewise analogous to what occurs in the supratentorial region, where both deep^4, 5^ and mixed^29^ ICH are driven mostly by hypertension.

Our findings provide external validation of previous studies showing that the location of cICH largely mirrors that of supratentorial brain regions. Furthermore, our study has several additional strengths. First, we used MRI markers of cSVD and the latest version of the Boston criteria 2.0 to affirm the association between CAA diagnosis and superficial cICH and further enhance the precision of the results. Second, our overall sample size and available MRI studies exceed those in previous similar studies.^20^ Our findings have implications for both research and clinical practice. They could be employed to assess patients’ eligibility for clinical trials targeting CAA and treatment decisions that may be location-dependent. However, our study has some limitations. The most significant is the lack of pathology samples to confirm a definite diagnosis of CAA based on the Boston criteria 2.0. Nevertheless, our findings are consistent with previous pathological^16, 18, 19^ and imaging^20^ evidence suggesting that CAA is the primary etiology of superficial cICH and HTN the one of deep cICH. Another limitation is that MRI was not obtained in all patients included in this analysis, which might have introduced bias. *Table S1* shows the differences in baseline characteristics between patients with cerebellar ICH with and without MRI during admission. Although the results of this single-center study may not be generalizable to other cICH populations, they are largely in line with previous research. Overall, our results are exploratory and should be considered hypothesis-generating for future studies.

## Conclusions

Our results suggest that topographic localization within the cerebellum might be associated with different underlying pathophysiologic processes, with superficial cICH more likely to be associated with CAA, whereas deep/mixed cICH is almost exclusively linked to hypertensive arteriopathy.

## Data Availability

The data will be available upon reasonable request.

